# Platelets can contain SARS-CoV-2 RNA and are hyperactivated in COVID-19

**DOI:** 10.1101/2020.06.23.20137596

**Authors:** Younes Zaid, Florian Puhm, Isabelle Allaeys, Abdallah Naya, Mounia Oudghiri, Loubna Khalki, Youness Limami, Nabil Zaid, Khalid Sadki, Rafiqua Ben El Haj, Wissal Maher, Belayachi Lamiae, Bouchra Belefquih, Amina Benouda, Amine Cheikh, Yahia Cherrah, Louis Flamand, Fadila Guessous, Eric Boilard

## Abstract

**Rationale:** In addition to the overwhelming lung inflammation that prevails in COVID-19, hypercoagulation and thrombosis contribute to the lethality of subjects infected with severe acute respiratory syndrome coronavirus 2 (SARS-CoV-2). Platelets are chiefly implicated in thrombosis. Moreover, they can interact with viruses and are an important source of inflammatory mediators. While a lower platelet count is associated with severity and mortality, little is known about platelet function during COVID-19.

**Objective:** To evaluate the contribution of platelets to inflammation and thrombosis in COVID-19 patients.

**Methods and Results:** We document the presence of SARS-CoV-2 RNA in platelets of COVID-19 patients. Exhaustive assessment of cytokines in plasma and in platelets revealed the modulation of platelet-associated cytokine levels in COVID-19, pointing to a direct contribution of platelets to the plasmatic cytokine load. Moreover, we demonstrate that platelets release their alpha- and dense-granule contents and phosphatidylserine-exposing extracellular vesicles. Functionally, platelets were hyperactivated in COVID-19 subjects, with aggregation occurring at suboptimal thrombin concentrations. Furthermore, platelets adhered more efficiently onto collagen-coated surfaces under flow conditions.

**Conclusions:** These data suggest that platelets could participate in the dissemination of SARS-CoV-2 and in the overwhelming thrombo-inflammation observed in COVID-19. Thus, blockade of platelet activation pathways may improve outcomes in this disease.

**KEY POINTS:** Platelets are a source of inflammatory cytokines and degranulate in COVID-19 Platelets contain SARS-CoV-2 RNA molecules and are prone to activation in COVID-19

**Subject terms:** Infectious diseases/Emerging infectious diseases, SARS-CoV-2, COVID-19, Hematology, Platelets

## Introduction

The outbreak of coronavirus disease 2019 (COVID-19), caused by severe acute respiratory syndrome coronavirus 2 (SARS-CoV-2) infection, emerged in Wuhan of Hubei province in China in December 2019 and rapidly spread to more than 196 countries worldwide.^1,2^ The World Health Organization (WHO) declared the outbreak a serious public health emergency of international concern and by June 2020, the virus had infected over 7 million and killed more than 400 000 individuals worldwide.^2,3^

SARS-CoV-2 is a positive-sense single stranded RNA virus.^4,5^ The main SARS-CoV-2 counter-receptor on human cells is the angiotensin converting enzyme 2 (ACE2), highly expressed by nasopharyngeal airway epithelial cells as well as alveolar epithelial cells, vascular endothelial cells and lung macrophages. The virus tropism likely explains the notorious respiratory symptoms observed during infection.^6,7^ An uncontrolled systemic inflammatory response, known as the cytokine storm, results from immune effector cell release of substantial amounts of pro-inflammatory cytokines, such as tumor necrosis factor (TNF), interleukin (IL)-1, IL6, IL7 and granulocyte-colony stimulating factor (G-CSF),^8,9^ which are suggested to contribute to SARS-CoV-2 lethality.^1,10,11^ While outstanding research aimed at understanding SARS-CoV-2 pathogenesis initially focused on lung inflammation, COVID-19 also implicates multi-organ damage, leading to multiple organ failure, notably of the respiratory, cardiac, renal and hepatic systems.^1,12^

Thrombotic complications that manifest as microvascular thrombosis, venous or arterial thrombosis are features detected in most patients with multi-organ failure.^13-17^ Disseminated intravascular coagulation (DIC) is a condition in which blood clots form throughout the body, blocking small blood vessels. Although DIC appeared rarely and was associated with bleeding manifestations in a multicenter retrospective study involving 400 COVID-19 patients (144 critically ill),^17^ DIC occurred in the majority (71%) of patients who died of COVID-19 according to a study of 183 consecutive patients.^18^ What drives DIC during COVID-19 is currently unknown. It is important to note that accumulating evidence shows that the blood of COVID-19 patients is hypercoagulable.^14,17-20^ Higher D-dimer and fibrin degradation product (FDP) levels, a longer prothrombin time and activated partial thromboplastin time are observed in SARS-CoV-2-infected patients with hospitalization complications or in those who died, relative to survivors.^17,18,20^

Hematological manifestation in symptomatic COVID-19 patients include severe leukopenia and lymphopenia.^14,18,21^ Lower platelet counts are associated with increased risk of in-hospital mortality in COVID-19 patients, although the platelet levels are generally not considered as clinically-relevant as patients typically do not require platelet transfusions.^17,22-26^ It is suggested that SARS-CoV-2 may reduce platelet production, increase platelet destruction, or more likely that platelet activation and thrombosis in patients may increase platelet consumption.^25^ As thrombosis and blood coagulation are chiefly controlled by platelets, it is critical to define whether platelets are activated in COVID-19.

Platelets are small (2–4 µm in diameter) anucleated cells derived from megakaryocytes in bone marrow and lungs.^27,28^ Nearly one trillion platelets patrol the blood vessels to maintain the integrity of the vasculature. Damage to blood vessels triggers the formation of a thrombus to stop subsequent bleeding.^29^ Thrombus formation mediated by platelets can also implicate extracellular vesicles (microparticles or microvesicles), which provide anionic phospholipids that can support the coagulation cascade.^30^

In addition to their role in hemostasis and thrombosis, platelets also contribute to immunity and inflammation.^31-33^ The extravasation of neutrophils and their invasion of inflamed tissues require their interaction with activated platelets.^34,35^ Moreover, the liberation of neutrophil-extracellular trap (NETosis), a process by which neutrophils release extracellular DNA, is observed in COVID-19.^36,37^ NETosis requires platelets and may thus contribute to thrombosis.^38,39^ Platelets express immune and inflammatory molecules such as interleukin-1 (IL1),^40^ and a set of immune receptors including CD40L, Toll-like receptors (TLR),^31^ and the Fc receptor for IgG FcγRIIA^41^.

Viruses such as dengue and influenza can infect megakaryocytes, which leads to the upregulation of type-I interferon genes.^42^ While dengue virus can replicate in platelets,^43^ influenza virus can be internalized by platelets and is transported through the circulatory system in humans.^44^ Influenza virus-induced lung injury in mice can be prevented by targeting of platelet αIIbβ3 by an antagonist or other anti-platelet compounds such as clopidogrel, an antagonist of ADP receptors, or blockers of protease-activated receptor-4, thus pointing to an active role by platelets in influenza pathogenesis.^45^ Influenza and herpes simplex virus-1 also activate platelet aggregation and thrombosis due to the presence of prevalent opsonizing antibodies and their interaction with FcγRIIA.^46,47^ Thus, activated platelets may contribute to both the overwhelming inflammation and thrombosis that prevail in COVID-19.

While thrombosis and coagulation abnormalities predict worse outcomes in COVID-19, the role of platelet function has yet to be investigated. Here, we examined platelet activation, secretion, adhesion and aggregation in patients with severe and non-severe COVID-19 infection.

## Methods

Supplement contains the detailed description of methods.

## Results

### Patient screening and characterization

A total of 1,544 patients with suspected SARS-CoV-2 infection (travelers, proximity to infected patients and presence of symptoms) were screened according to the flow chart presented in **Online Figure I**. The clinical symptoms included fever, cough, dyspnea, fatigue, headache, chest pain and pharyngalgia. Initial screening in addition to laboratory blood testing and high-resolution chest computed tomography imaging (CT scans) (**Online Figure II**) led to the selection of 186 patients, who were then screened for the presence of 2019-nCoV RdRp and E genes using RT-PCR (**Online Figure III**) (see **Table 1 and Online Figure IV** for clinical data). The degree of COVID-19 severity (severe *vs* non-severe) was defined at the time of patient admission using the American Thoracic Society guidelines for community-acquired pneumonia ^48^. There were 71 cases in the non-severe COVID-19 (COVID-Non-Severe) group and 34 cases in the severe COVID-19 group (COVID-Severe), while 81 individuals were declared negative by both throat swab RT-PCR amplification of 2019-nCoV RdRp and E genes and chest CT scans. Among all COVID-19 positive patients, the mean ages were 48.96 ± 17.96 and 57.15 ± 23.10 years for non-severe cases and severe cases, respectively. The hospitalization duration was significantly higher for severe patients (29.63 ± 2.74 days) compared with non-severe patients 18.27 ± 4.33 (days), with 8.5% of total patients critically ill and admitted to the Intensive Care Unit (ICU). Four COVID-Severe patient deaths occurred in the ICU, while all non-severe patients survived.

**Table 1.**
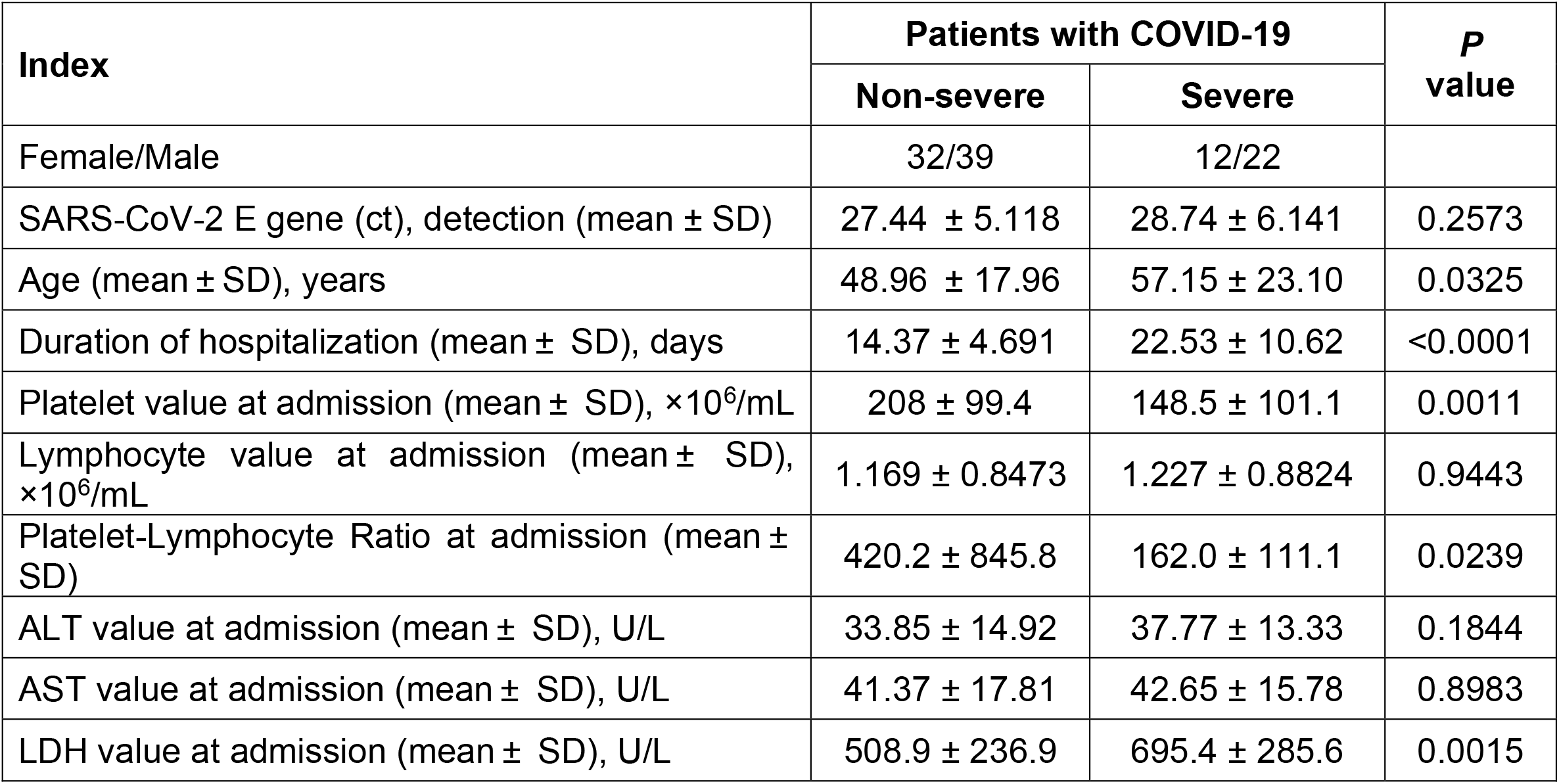
Clinical Analysis and Blood Parameters. ALT = Alanine Transaminase; AST = Aspartate Transaminase; LDH = Lactate Dehydrogenase

| Index | Patients with COVID-19 |  | *P*<br>value |
| --- | --- | --- | --- |
|  | Non-severe | Severe |  |
| Female/Male | 32/39 | 12/22 |  |
| SARS-CoV-2 E gene (ct), detection (mean ± SD) | 27.44 ± 5.118 | 28.74 ± 6.141 | 0.2573 |
| Age (mean ± SD), years | 48.96 ± 17.96 | 57.15 ± 23.10 | 0.0325 |
| Duration of hospitalization (mean ± SD), days | 14.37 ± 4.691 | 22.53 ± 10.62 | <0.0001 |
| Platelet value at admission (mean ± SD), $\times 10^6/\text{mL}$ | 208 ± 99.4 | 148.5 ± 101.1 | 0.0011 |
| Lymphocyte value at admission (mean ± SD), $\times 10^6/\text{mL}$ | 1.169 ± 0.8473 | 1.227 ± 0.8824 | 0.9443 |
| Platelet-Lymphocyte Ratio at admission (mean ± SD) | 420.2 ± 845.8 | 162.0 ± 111.1 | 0.0239 |
| ALT value at admission (mean ± SD), U/L | 33.85 ± 14.92 | 37.77 ± 13.33 | 0.1844 |
| AST value at admission (mean ± SD), U/L | 41.37 ± 17.81 | 42.65 ± 15.78 | 0.8983 |
| LDH value at admission (mean ± SD), U/L | 508.9 ± 236.9 | 695.4 ± 285.6 | 0.0015 |

### Blood parameters and thrombocytopenia

The severity of disease was established, and routine blood parameters were monitored at the time of admission for both the non-severe and severe cases of COVID-19 (**Table 1 and Online Figure IV**).^2^ Lymphocyte counts, alanine aminotransferase (ALT) and aspartate aminotransferase (AST) data were similar between the 2 groups of infected patients. However, the lactate dehydrogenase (LDH) inflammation marker, which was higher than the expected normal range in both groups, was significantly increased in severe COVID-19 patients in comparison with non-severe patients, pointing to a possible cytokine storm associated with disease severity (**Table 1 and Online Figure IV)**.

Thus, we evaluated platelet numbers and platelet-to-lymphocyte ratios (PLR) measured at the time of admission in the two groups. Platelet counts were in the lower range in both COVID-19 severe (148.5 ± 10.1) and non-severe patients (208.0 ± 99.4) in comparison to the expected count in healthy individuals (130 – 400 × 10^6^/mL). Moreover, we observed a modest, but statistically significant reduction in platelet counts between the 2 patient groups. The PLR was lower in severe patients compared with non-severe patients, arguing in favor of the thrombosis mainly reported in more severe cases (**Table 1 and Online Figure IV**).

### Platelets in COVID-19 patients can be associated with SARS-CoV-2 RNA

SARS-CoV-2 RNA has been identified in urine and stool samples in addition to semen, consistent with expression of ACE2 outside the respiratory tract, such as in endothelial cells that line blood vessels, heart, kidney and spermatozoa.^51-54^ Whether platelets express ACE2 in COVID-19 and contain SARS-CoV-2 RNA have never been verified. Thus, platelets were isolated from recruited healthy individuals (n=17) and from COVID-19 patients (38 non-severe and 11 severe) at the time of admission, and we tested for the presence of RNA coding for SARS-CoV-2 RdRp and E, and for ACE2. While no RNA from SARS-CoV-2 was detected in healthy subjects (0%), we detected SARS-CoV-2 RNA in platelets from non-severe (23.7%) and severe (18.2%) COVID-19 patients. Both SARS-CoV-2 RdRp and E genes were amplified equally (**Table 2 and Online Figure VA**), which might suggest that platelets contain viral particles. Conversely, ACE2 mRNA molecules were identified in platelets of all individuals tested (10 individuals/group) (**Table 2 and Online Figure VB**). The data suggest that platelets may disseminate SARS-CoV-2 RNA molecules.

**Table 2.** Detection of SARS-CoV-2 and ACE2 (mRNA) in platelets. ACE2 = Angiotensin-converting enzyme 2; ct = threshold cycle

| Index |  |  | Patients with COVID-19 |  |
| --- | --- | --- | --- | --- |
|  |  | Healthy | Non-Severe | Severe |
| **SARS-CoV-2**<br>**E gene** | detection in platelets<br>(positive/total donors) | 0/17 | 9/38 | 2/11 |
| | ct values (mean ± SD) | n.d. | 28.89 ± 5.86 | 34 ± 2.82 |
| **ACE2, mRNA** | detection in platelets<br>(positive/total donors) | 10/10 | 10/10 | 10/10 |
| | ct values (mean ± SD) | 27.73 ± 1.41 | 26.43 ± 0.53 | 26.93 ± 0.34 |

### Circulating cytokines

Elevated inflammatory cytokine levels are reported in the blood of COVID-19 patients^8,9^ and may contribute to the overwhelming inflammation and to coagulation through the activation of endothelial cells.^55,56^ We used a multiplex assay to monitor 48 cytokines in plasma prepared from healthy individuals (n=10) and from COVID-19 patients (10 non-severe and 9 severe) at the time of admission. With the exception of 12 cytokines/chemokines that were unchanged (IL-1α, IL-4, IL-5, IL-8, IL-10, IL-12p70, IL-15, MIP1 α, MCP-1, CXCL9, IL-25, RANTES), and IL-3 the concentration of which was significantly reduced in COVID-19 (non-severe and severe), we identified 35 cytokines that were significantly elevated, including TNF, IL-1, IL-6, interferon (IFN) α and γ, and eotaxin (**Figure 1 and Online Figure VI**). Soluble CD40L, as well as growth factors such as granulocyte colony-stimulating factor (G-CSF), granulocyte-macrophage colony-stimulating factor (GM-CSF), fibroblast-growth factor 2 (FGF-2), platelet-derived growth factor (PDGF) and vascular endothelial growth factors (VEGF) were also significantly increased in COVID-19 (**Figure 1 and Online Figure VI**).

**Figure 1.**
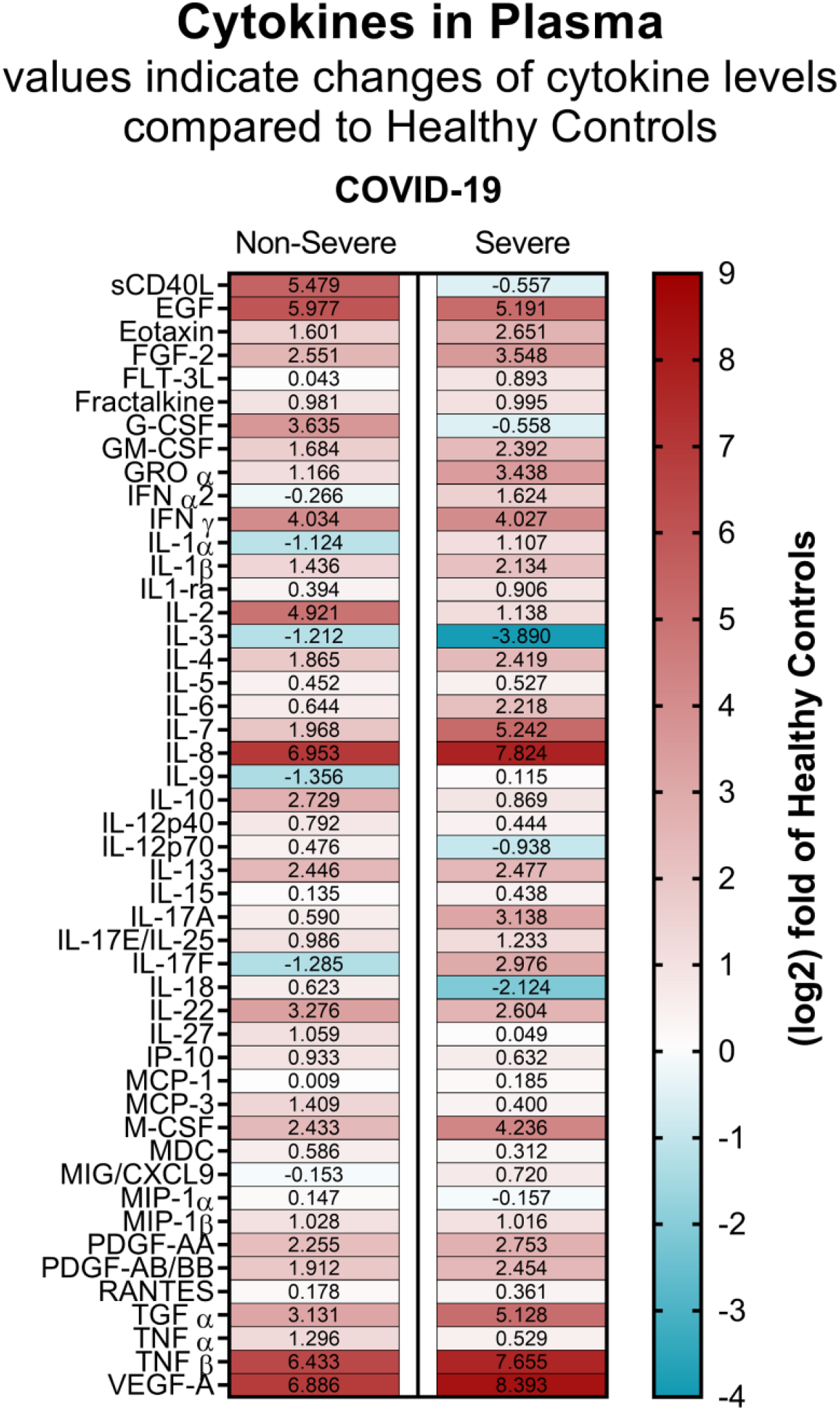
Cytokine and chemokine levels in plasma from COVID-19 patients. Heat map visualization of 48 cytokine/chemokine expression profiles in plasma of COVID-19 non-severe (mean of n=10) and severe (mean of n=9) patients relatively to the heathy controls (mean of n=10). Cytokine/chemokine expression is represented as a (log2) fold change relative to healthy controls. The numbers in each panel represent the mean change and the color codes refer to red for increased expression and blue for decreased expression. Absolute cytokine/chemokine values (pg/mL) and statistical analysis are shown in **Online Figure VI**.

### Platelets produce inflammatory cytokines in COVID-19

We then determined the ability of COVID-19 patient platelets to produce inflammatory mediators. Platelets derived from healthy individuals and from severely and non-severely affected patients were stimulated with low doses of α-thrombin (0.025 and 0.05 U/mL). Platelets were then lysed, and the amounts of IL-1β, IL-18, sCD40L and the eicosanoid thromboxane B_2_ (TxB_2_) were determined. While IL-18 secretion and thromboxane production were unaffected in the disease, we found that COVID-19 platelets were more potent at producing IL-1β and soluble CD40L upon exposure to 0.05 U/mL of thrombin in comparison with healthy subjects (**Figure 2A**). Platelets from all subjects were similarly efficient at releasing the inflammatory mediators when higher concentrations of thrombin were used, suggesting that platelets are a source of inflammatory mediators and may be primed to release certain inflammatory molecules during SARS-CoV-2 infection.

**Figure 2.**
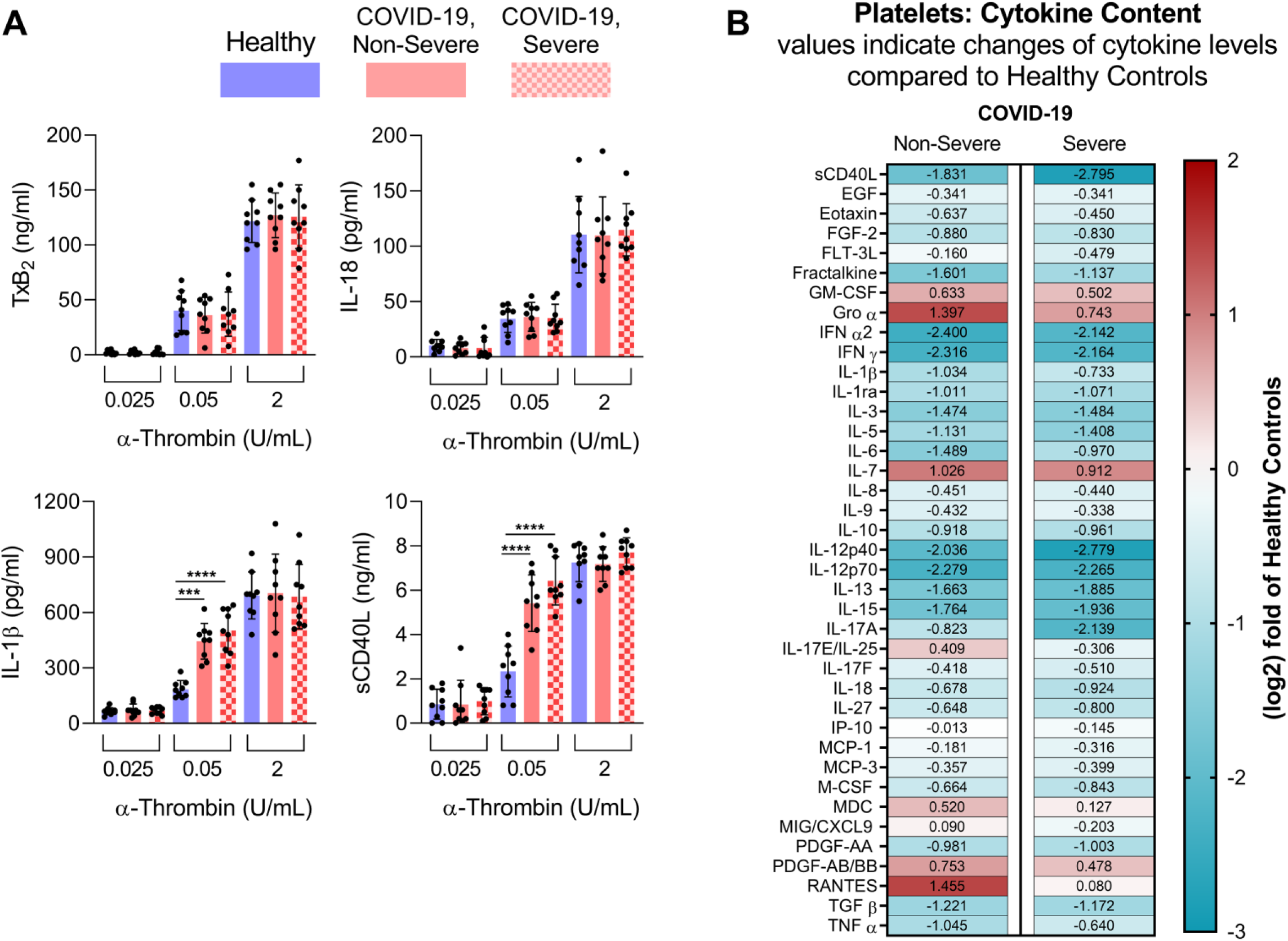
Platelets are prone to produce and release inflammatory molecules in COVID-19 patients. **A**) Platelets from healthy controls (n=9), COVID-19 non-severe (n=9) and COVID-19 severe (n=9) patients were stimulated with 0.025, 0.05 or 2 U/mL of α-thrombin. Thromboxane B2 (TxB2), interleukin (IL)-18, IL-1β and soluble CD40 ligand (sCD40L) production was evaluated. Data are represented as mean ± SD. Statistical analysis: one-way ANOVA. ***P<0.001 and ****P<0.0001. **B**) Heat map visualization of 39 cytokine/chemokine expression profiles in platelet content of COVID-19 non-severe (mean of n=10) or severe (mean of n=9) patients compared to the heathy controls (mean of n=9). Cytokine/chemokine expression is represented as a (log2) fold change relative to healthy controls. The numbers in each panel represent the mean change and the color codes refer to red for increase expression and blue for decrease expression. Absolute cytokine/chemokine values (pg/mL) and statistical analysis are shown in **Online Figure VI**.

Because they are anucleated, the platelet cytokine content can mirror that of megakaryocytes, or may be regulated by translation upon platelet activation.^57^ However, nothing is known regarding the platelet cytokine content in COVID-19. Thus, we monitored cytokines in lysates prepared using platelets from COVID-19 patients and included healthy subjects as controls. While a few cytokines were below the detection limit (G-CSF, IL-1α, IL-2, IL-4, IL-5, IL-6, IL-10, IL-22, TGF α and VEGF), we detected 36 cytokines in platelets from healthy individuals, including CD40L, IFN α and γ and IL-1β (**Figure 2B and Online Figure VI**), in agreement with their important cytokine content. Of note, we found significantly reduced levels for 17 cytokines, including cytokines relevant to virus and inflammatory responses (e.g. sCD40L, eotaxin, IFN α and γ, IL1β, TNF α and β) (**Figure 2B and Online Figure VI**) in COVID-19. Together, the data show that platelets are primed to generate certain cytokines and release their cytokine cargo during SARS-CoV-2 infection.

### Platelets degranulate in COVID-19

Platelet alpha- and dense-granule content, determined by the assessment of PF4 and serotonin respectively, was examined in platelets. We found a reduction in both PF4 and serotonin in platelets from non-severe and severe COVID-19 patients in comparison with healthy individuals (**Figure 3A**). Moreover, robust increases in PF4 and serotonin levels were measured in plasma from COVID-19 patients, independent of disease severity (**Figure 3B**), thereby suggesting that platelets degranulate during SARS-CoV-2 infection.

**Figure 3.**
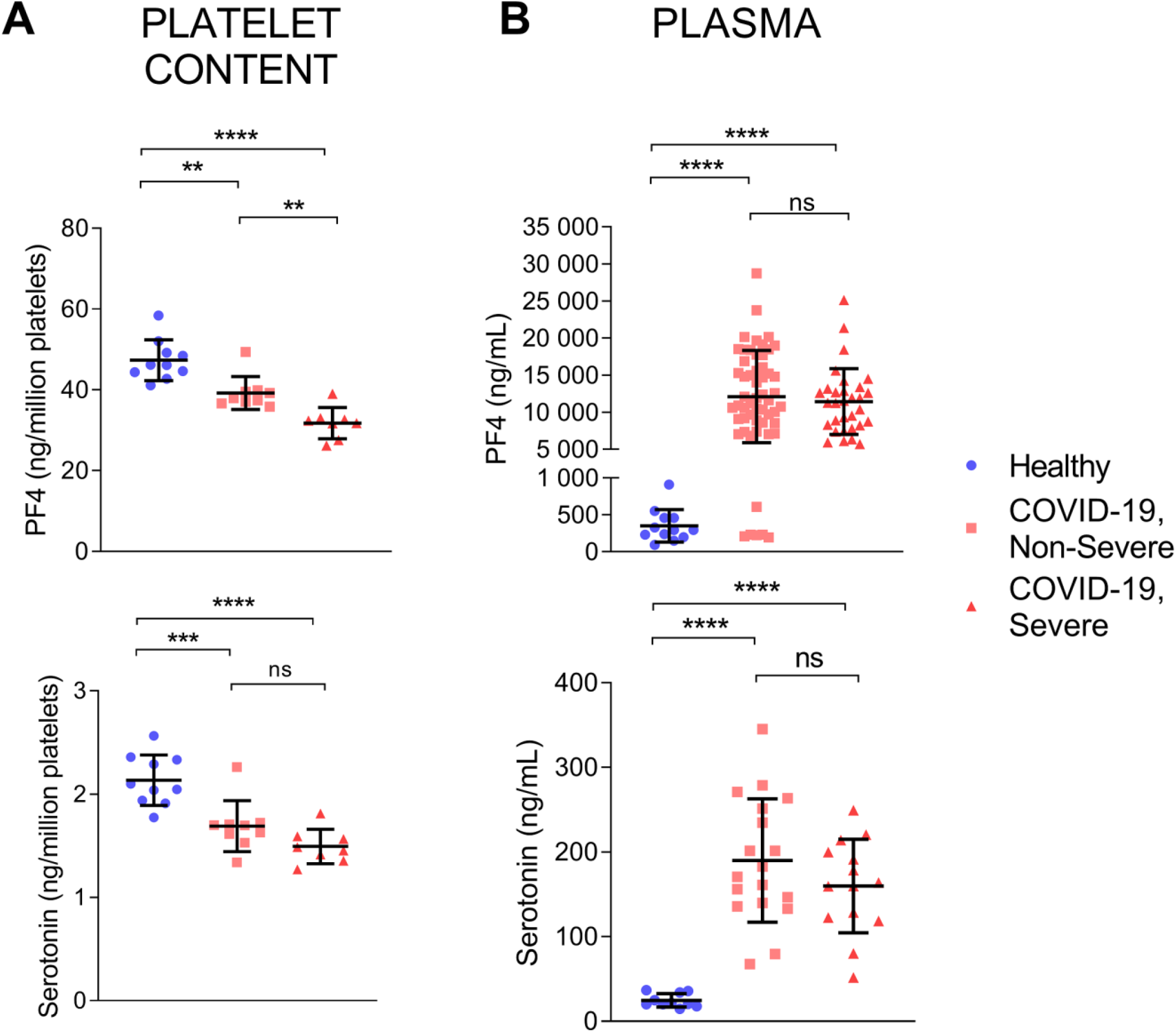
Platelets are degranulated in COVID-19 patients. Markers of platelet degranulation (PF4 for alpha granules and serotonin for dense granules) were evaluated in plasma of COVID-19 patients. Concentrations of PF4 (upper panels) and serotonin (lower panels) were measured in platelets (**A**) and plasma (**B**) from healthy controls, COVID-19 non-severe and COVID-19 severe patients. For platelet content, values were expressed as ng per million platelets. Data are represented as mean ± SD. Statistical analysis: ROUT method identified two outliers for serotonin (platelet content), which were thus excluded from the analysis. one-way ANOVA. **P<0.01,***P<0.001 ****P<0.0001, ns (non-significant). PF4: healthy controls (n=12 for plasma, n=10 for platelet content), COVID-19 non-severe (n=55 for plasma, n=10 for platelet content), COVID-19 severe (plasma n=31, platelet content n=9). Serotonin: healthy controls (n=10 for plasma, n=10 for platelet content), COVID-19 non-severe (n=18 for plasma, n=9 for platelet content), COVID-19 severe (plasma n=14, platelet content n=8).

### Platelets release extracellular vesicles in COVID-19

Extracellular vesicles (EV) released by platelets can participate in both inflammation and the coagulation process due to the exposure of phosphatidylserine^30,58,59^. However, it is currently unknown whether EV are produced in COVID-19. EV smaller than 1 µm and of platelet origin identified by the surface expression of CD41 were detected by high sensitivity flow cytometry. To determine whether EV expressed surface phosphatidylserine, we included annexin V conjugated with a fluorescent probe in the analyses. EV were sensitive to detergent treatment, which confirmed their membrane moiety. Moreover, no annexin V^+^ EV were detected when Ca^2+^ ions were chelated by ethylenediaminetetraacetic acid (EDTA), which further ensured the specificity of the detection approach (**Online Figure VII**). We found a statistically significant increase in platelet-derived EV and in phosphatidylserine-exposing platelet EV in COVID-19 patients in comparison with healthy subjects (**Figure 4**). The levels of both types of EV were similar in non-severe and severe COVID-19 patients.

**Figure 4.**
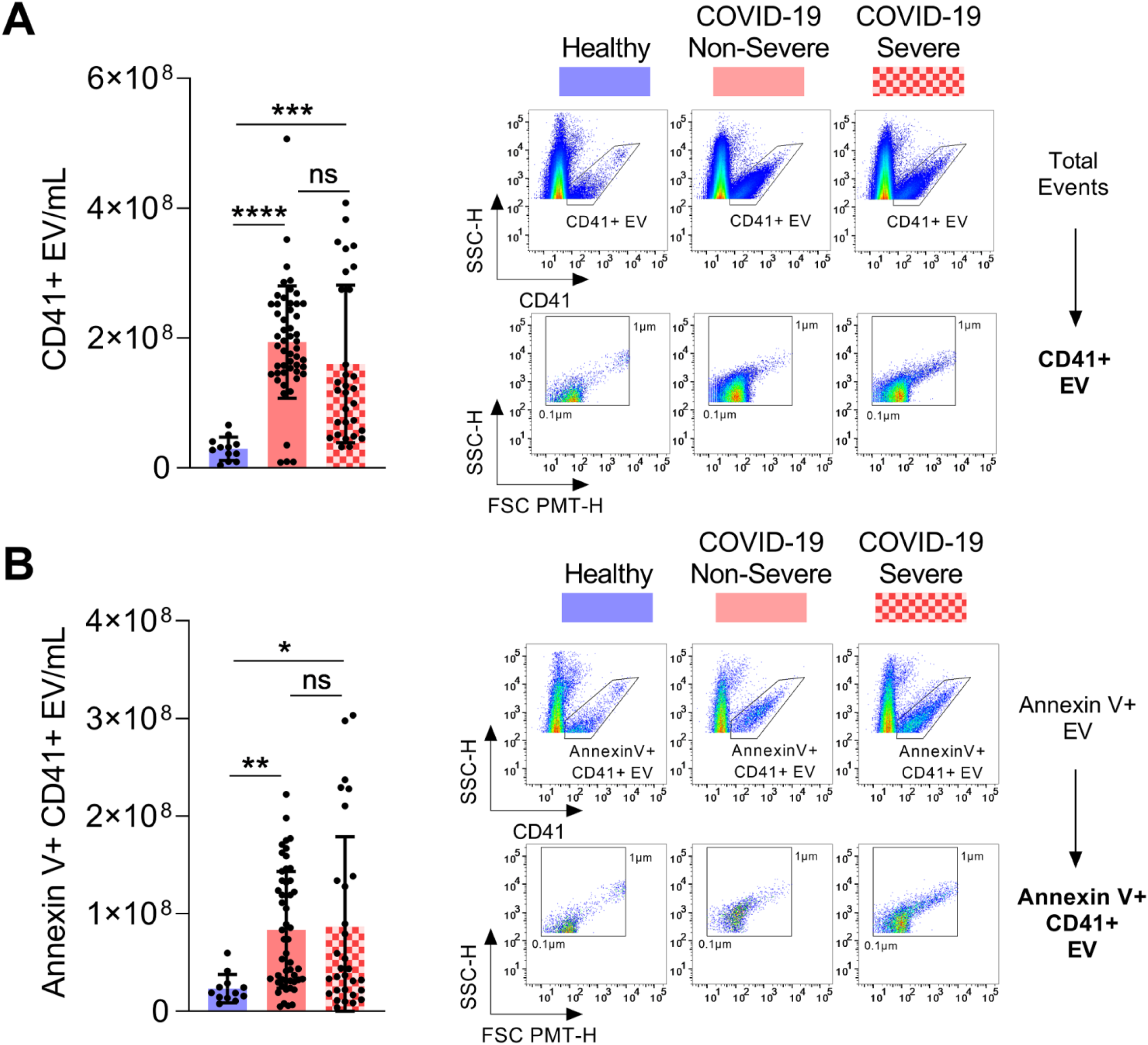
Platelet extracellular vesicles are released in COVID-19 patients. Circulating platelet extracellular vesicles (CD41+ EV) expressing phosphatidylserine or not were analyzed in plasma from healthy controls (n=12), COVID-19 non-severe (n=51) and COVID-19 severe patients (n=30). **A**) Total CD41+ EV were quantified (left panel) and representative scatter plots of CD41+ EV relative size and inner complexity (FSC=forward scatter, SSC=sideward scatter) are illustrated (right panels). **B**) Annexin V+ CD41+ EV were quantified (left panel) and representative scatter plots of AnV+CD41+ EV relative size are illustrated (right panels). The gating strategy is illustrated in **Online Figure VII**. Data are represented as mean ± SD. Statistical analysis: Data were not normally distributed (Shapiro-Wilk test). Kruskal-Wallis test *P<0.05, **P<0.01, ***P<0.001 and ****P<0.0001.

### Platelets are hyperactivated in COVID-19

Platelet activation is a complex balance of positive and negative signaling pathways involving several protein kinase C (PKC) isoforms expressed in human platelets. Among these kinases, PKCδ is a key regulator of platelet granule secretion, activation and aggregation activity.^60,61^ To determine whether PKCδ is involved in platelet activation during this infection, platelets collected from severe and non-severe patients were stimulated (or not) with a low dose of α-thrombin (0.05 U/mL), lysed and then analyzed for the presence of phosphorylated-PKCδ Tyr^311^. We found that PKCδ phosphorylation on Tyr^311^ residue was significantly increased in response to a low dose of α-thrombin in patients with severe (p < 0.01) and non-severe (p < 0.05) COVID-19 infection, while phosphorylation was undetectable in controls. Phosphorylation of PKCδ was also significantly increased in severe patients in comparison with non-severe patients (**Figure 5**), thus suggesting that platelet activation signaling pathways may be upregulated in the disease.

**Figure 5.**
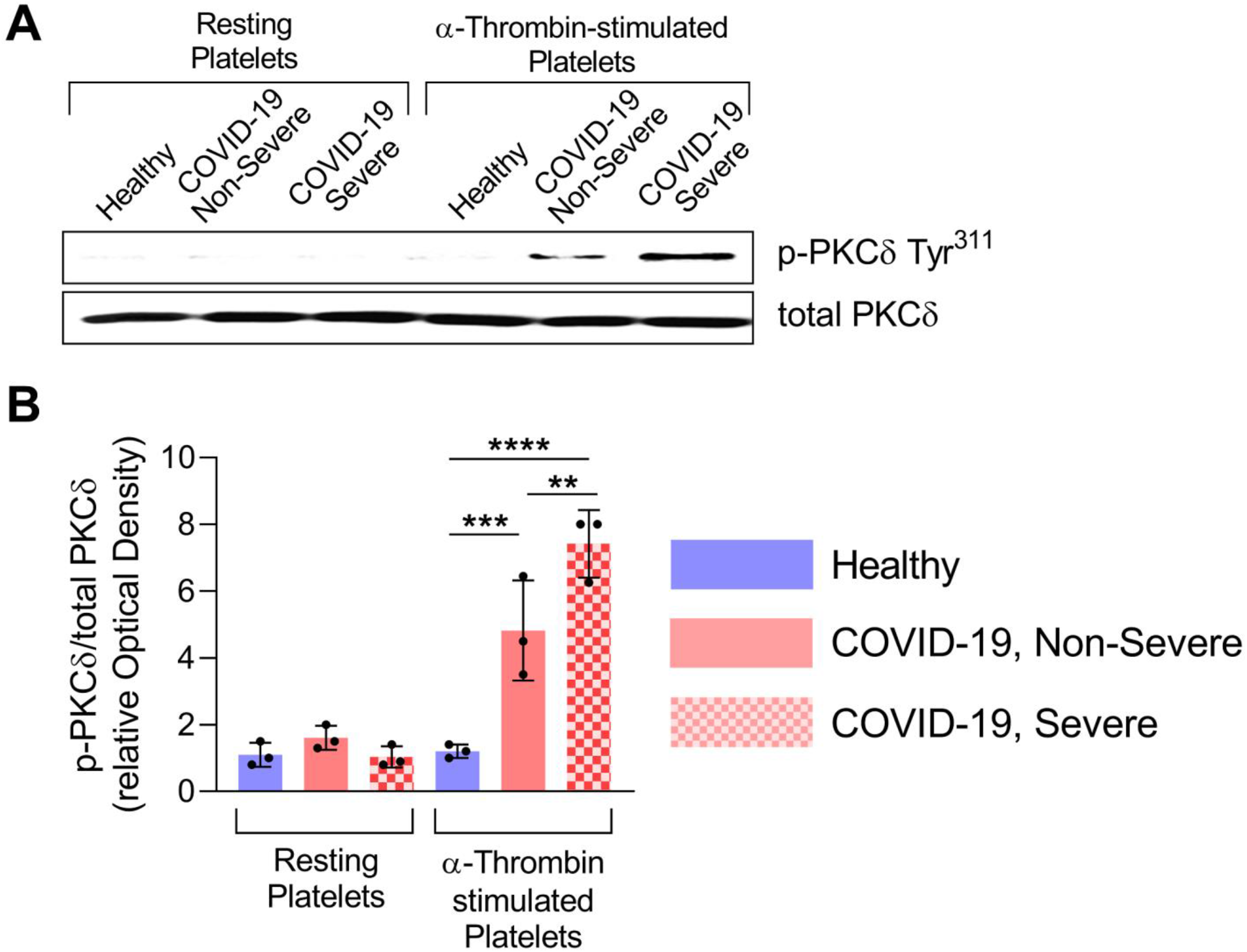
PKCδ phosphorylation is increased in platelets of COVID-19 patients. PKCδ phosphorylation on Tyr311 residue is increased in response to α-thrombin in patients with severe and non-severe COVID-19 infection. Platelets were stimulated (or not) with 0.05 U/mL of α-thrombin for 5 min. Platelet lysates were analyzed by SDS-PAGE for p-PKCδ Tyr311. Total PKCδ was evaluated in each condition. **A**) Immunoblot representative of 3 donors. **B**) Densitometric analysis of p-PKCδ Tyr311 normalized to total PKCδ was performed, data were expressed as relative Optical Density (n=3). Data are represented as mean ± SD. Statistical analysis: one-way ANOVA. **P<0.01, ***P<0.001 and ****P<0.0001.

The relationship between platelet activation and the severity of COVID-19 was then investigated by functional assays. Platelets from severe and non-severe COVID-19 patients were collected and analyzed by optical aggregometry under arterial flow conditions using various doses of α-thrombin. Platelets exposed to suboptimal concentrations of α-thrombin (0.05 U/mL) were highly activated (≥2.5-fold increase) in both severe and non-severe patients compared with controls **(Figure 6A and 6B)**. When platelets from all groups were stimulated with a higher concentration of α- thrombin (2 U/mL), no differences were observed, pointing to a lower platelet stimulation threshold in COVID-19 patients (**Figure 6A and 6B**). Moreover, the formation of platelet aggregates was visualized by rhodamine-based immunofluorescence on a collagen-coated surface under flow conditions, and the ratio of adherent platelets to total platelets (activated vs non-activated platelets) was calculated. The number of adherent platelets was significantly higher in severe patients compared with non-severe patients and controls, implying the involvement of the thrombotic events in the disease magnitude (**Figure 6C**).

**Figure 6.**
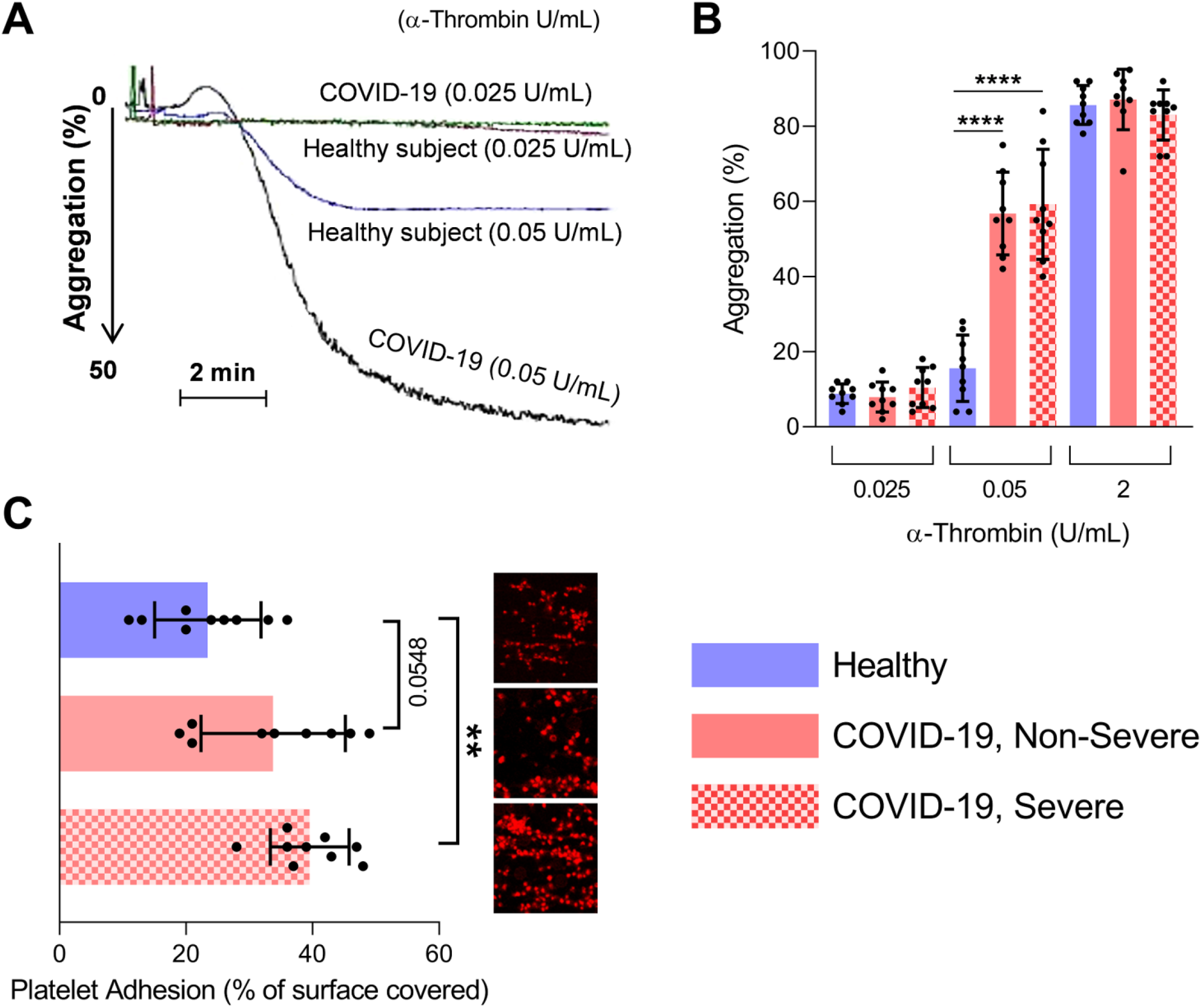
Platelets are prone to aggregation in COVID-19 patients. Platelet aggregation and adhesion were evaluated in healthy donors, COVID-19 non-severe and COVID-19 severe patients (n=9). **A**), **B**) Platelets were stimulated with 0.025, 0.05 or 2 U/mL. **A**) Representation of light transmission curve of platelets aggregation from healthy controls and COVID-19 patients. **B**) Quantification (%) of platelet aggregation in healthy controls (n=9), COVID-19 non-severe (n=9) or COVID-19 severe (n=9) patients. **C**) Platelet adhesion on collagen was evaluated under flow condition after 5 min. Data are presented as percentage of surface covered by platelets. A representative image for each condition is illustrated. Data are represented as mean ± SD. Statistical analysis: one-way ANOVA. **P<0.001.

The results are visualized in a graphical summary in **Figure 7**.

**Figure 7.**
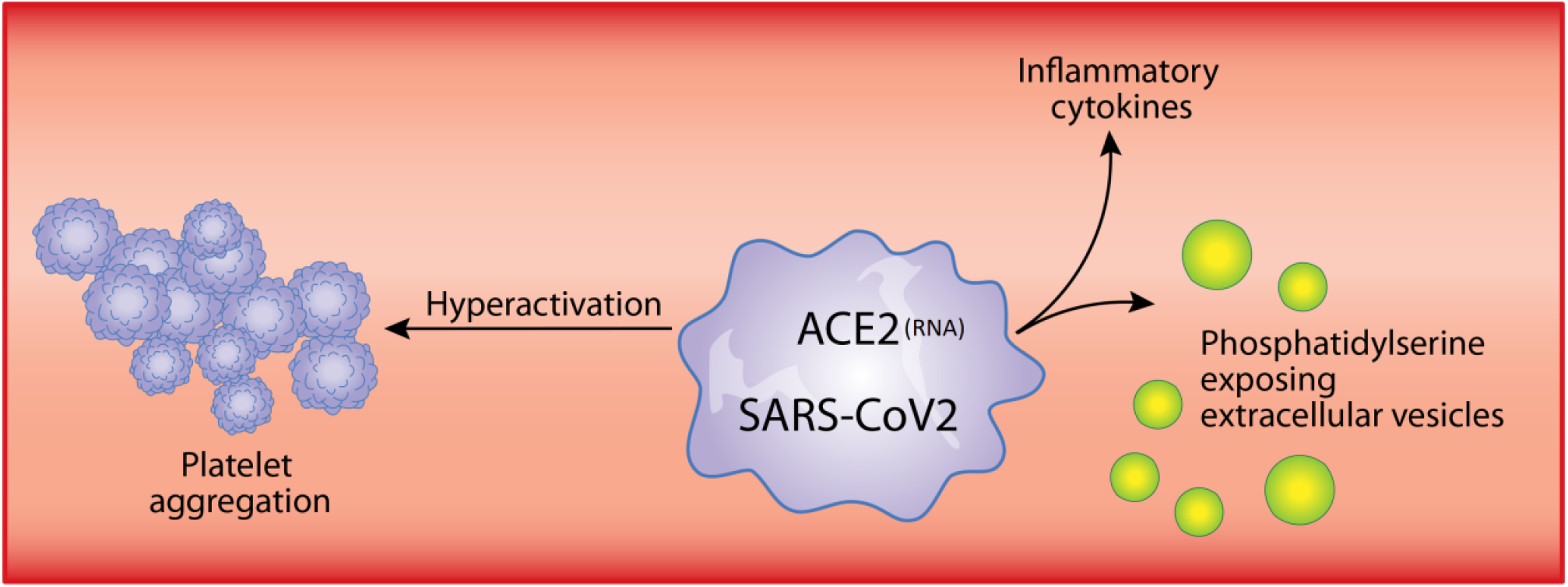
Graphical Summary. **Novelty and Significance** a. What is known? In addition to the overwhelming inflammation that prevails in COVID-19, hypercoagulation and thrombosis are now recognized hallmark of severe COVID-19 and contribute to the lethality of severe acute respiratory syndrome coronavirus 2 (SARS-CoV-2). Platelets are chiefly implicated in thrombosis and can interact with different microbes including viruses. Moreover, platelets are a major source of inflammatory mediators. b. What new information does this article contribute? This article documents that human platelets, in some patients (approximately 20%), contain SARS-CoV-2 RNA molecules. Moreover, platelets are hyperactivated in COVID-19: they have enhanced adhesion properties, degranulate (alpha and dense granule), and are a source of inflammatory cytokines and of extracellular vesicles. The data suggest that platelets could participate in the dissemination of SARS-CoV-2 and in the overwhelming thrombo-inflammation observed in COVID-19. Thus, blockade of platelet activation pathways may improve outcomes in this disease.

## Discussion

Both inflammation and thrombosis are clinical manifestations observed during SARS-CoV-2 infection. They can be lethal, and the understanding of cellular and molecular effectors in COVID-19 may reveal novel therapeutic approaches, critical for the treatment of patients whilst vaccines are still lacking. Platelets can interact with viruses and can participate in both inflammation and thrombosis. Thus, for this study, we examined platelets in a cohort of non-severe and severe COVID-19 patients with confirmed lower range (but not thrombocytopenic) platelet counts, in agreement with the current literature.^22-26^ Our findings identify SARS-CoV-2 RNA in patient platelets and reveal that platelets express pro-inflammatory molecules and are hyperactivated in COVID-19.

Spike glycoprotein S mediates SARS-CoV-2 attachment to the cellular receptor ACE2 and entry by fusion of the viral envelope with cell membranes.^62^ We detected ACE2 mRNA in platelets, which may contribute to the potential entry of the virus in platelets if translated and expressed. Of interest, was the detection of SARS-CoV-2 RNA in platelets from some COVID-19 patients. However, it remains to be established whether platelets represent target cells susceptible to infection or whether platelets might have captured circulating SARS-CoV-2 RNA molecules. Alternatively, megakaryocytes that produce platelets, may be susceptible to SARS-CoV-2 infection and transfer viral RNA as pro-platelets are produced. The presence of viral RNA in platelet cytosol may activate platelet TLR-7, a process that occurs in cases of influenza and encephalomyocarditis virus infections.^44,63^

Influenza virus and herpes simplex virus-1 (HSV-1) can trigger platelet degranulation when antibodies opsonize viral particles, and thereby activate platelet FcyRIIA.^46,47^ While antibodies specifically directed at SARS-CoV-2 are not expected in most individuals at an early stage of the pathogenesis, they might be produced at a later stage of infection, or prevailing cross-reacting antibodies against other more common coronaviruses that generate minor cold symptoms in humans (229E, NL63, OC43 and HKU1) may suffice to form immune complexes and activate platelet FcyRIIA.^4,5^ Thus, differences in antibody profiles, especially in neutralizing antibodies, among severe and non-severe COVID-19 patients may contribute to platelet interaction with the virus, and to the overwhelming inflammation. With the dissemination of the SARS-CoV-2 virus in the population, possible re-infections and vaccination, the activation of platelet FcyRIIA due to the presence of SARS-CoV-2 antibodies may be even more relevant in subsequent years.

SARS-CoV-2 entry into endothelial cells, which express ACE2, and the loss of endothelium integrity may favor recruitment of circulating platelets to sites of infection, their activation and degranulation, suggesting that the inhibition of endothelial cell activation may improve disease outcome.^64^ Megakaryocytes in lungs bear an immune profile distinct from the megakaryocytes found in bone marrow.^28^ While it is unknown if lung megakaryocytes express ACE2 protein, lung megakaryocytes may be infected, thereby leading to generation of virus-containing platelets in lungs. Moreover, dengue and influenza viruses can infect megakaryocytes, which impacts the megakaryocyte transcriptome.^42^ Hence, platelets produced by infected megakaryocytes, and/or by megakaryocytes in the inflamed environment, may present a different transcriptome, which may explain their potency at producing inflammatory cytokines, such as IL-1, when activated by α-thrombin.

From platelet degranulation and cytokine release analyses, we concluded that platelets can contribute to the cytokine storm reported in COVID-19. Only a limited number of cytokines (Gro α, IL-7, IL-12p40, PDGF AA/BB) were found significantly increased in platelets in COVID-19, which may be due to their over-expression by platelets or megakaryocytes, or to their abundance in the environment and their capture by platelets. While molecules such as CD40L, PF4 and serotonin are abundant in platelet granules and are released upon degranulation, cytokines such as IL-1 may be produced *de novo* following platelet activation.^57^ Therefore, the presence of CD40L, PF4 and serotonin are clear indicators of platelet degranulation *in vivo*. The molecules secreted by platelets can have effects on numerous cell types as well as the vasculature. Serotonin for instance, which we found to be abundantly released in this disease, can directly impact blood vessel integrity and promote leukocyte recruitment and cytokine release. ^46,65^ Of potential relevance, soluble CD40L, which we found to be released by platelets in COVID-19, can contribute to both activation of CD40 bearing cells and thrombosis by stabilizing glycoprotein αIIbβ3.^66^ This may not be unique to SARS-CoV-2 as dengue virus also induces the release of CD40L from human platelets.^67^

We made the original observation that platelets release extracellular vesicles (EV) in this disease. While EV, with their expression of phosphatidylserine can contribute to coagulation and thrombosis, they can also transport molecules from the mother platelets.^58^ EV could transport platelet-derived cytokines, as well as other pro-inflammatory molecules such as damage associated molecular patterns (DAMPs) reportedly transported by EV from activated platelets.^58,68,69^ The fact that platelets are activated in both non-severe and severe patients may explain the lower platelet counts observed in both cohorts of COVID-19 patients. As platelet degranulation and the release of EV were similar in both non-severe and severe COVID-19 patients, the contribution of other players that lead to severe manifestations of the disease is expected. Thus, therapeutic approaches to treatment of COVID-19 may require a combination of drugs targeting platelet activities or platelet-derived molecules in addition to medications directed against other inflammatory sources.

Using functional assays, we observed that platelets from COVID-19 patients are activated and hyperresponsive. Platelets were more efficiently at producing inflammatory cytokines, and they aggregated and adhered to a collagen surface more efficiently when originating from COVID-19 patients. As such, hyperactivation of platelets may contribute to the disease pathogenesis through both the release of inflammatory mediators and thrombosis. This could explain the lower platelet count and importantly, may contribute to the thrombo-inflammatory state associated with COVID-19 infection. In summary, our study has revealed that SARS-CoV-2 infection causes platelet activation resulting in increased systemic inflammatory mediator production and platelet aggregation, two main clinical symptoms associated with COVID-19. The blockade of platelet activation pathways may improve the outcomes in this disease.

## Data Availability

All data, analytic methods and study materials supporting the findings of this study are provided in the manuscript, supplemental material and available from the corresponding author upon reasonable request.

## Non-standard Abbreviations and Acronyms

ACE2: Angiotensin Converting Enzyme 2
ALT: Aminotransferase
AST: Aspartate Aminotransferase
COVID-19: Coronaviruse Disease 2019
FDP: Fibrin Degradation Product
IL: Interleukin
LDH: Lactate Dehydrogenase
NET: Neutrophil-extracellular trap
PF4: Platelet Factor 4
PKC: Protein kinase C
PLR: Platelet-Lymphocyte Ratio
SARS-CoV-2: Severe Acute Respiratory Syndrome Coronavirus 2
TLR: Toll-like receptor
TNF: Tumor Necrosis Factor
TxB2: Thromboxane B2

## Acknowledgments

We are grateful to the patients and the members of the COVID-19 laboratory of Cheikh Zaid Hospital in Rabat for their technical assistance with the RT-PCR tests, laboratory blood tests and the patients’ clinical data collection. YZ, FG and EB conceived the initial concept and designed the study. FP, IA, AN, MO, LK, YL, NZ, KS, RBE, BB, AC and YC participated to design the study, performed experiments and were involved in the data extraction. WM, LB and AB contributed critical reagents, biospecimens and instruments. EB, YZ, FP, IA, LF and FG wrote the manuscript and all authors read and approved the final manuscript.

## Ethics Statement

This study was approved by the Local Ethics Committee of Cheikh Zaid Hospital, Rabat, Morocco.

## Supplemental Materials

Expanded Materials & Methods Online Figures I – VII References 2, 48-50

## References

1. Tay MZ, Poh CM, Renia L, MacAry PA, Ng LFP. The trinity of COVID-19: immunity, inflammation and intervention. Nat Rev Immunol. 2020.

2. Guan WJ, Ni ZY, Hu Y, et al. Clinical Characteristics of Coronavirus Disease 2019 in China. N Engl J Med. 2020;382(18):1708–1720.

3. https://covid19.who.int CdC-WhOpoM. 2020.

4. Fehr AR, Perlman S. Coronaviruses: an overview of their replication and pathogenesis. Methods Mol Biol. 2015;1282:1–23.

5. Coronaviridae Study Group of the International Committee on Taxonomy of V. The species Severe acute respiratory syndrome-related coronavirus: classifying 2019-nCoV and naming it SARS-CoV-2. Nat Microbiol. 2020;5(4):536–544.

6. Xu H, Zhong L, Deng J, et al. High expression of ACE2 receptor of 2019-nCoV on the epithelial cells of oral mucosa. Int J Oral Sci. 2020;12(1):8.

7. Kuba K, Imai Y, Rao S, et al. A crucial role of angiotensin converting enzyme 2 (ACE2) in SARS coronavirus-induced lung injury. Nat Med. 2005;11(8):875–879.

8. Huang C, Wang Y, Li X, et al. Clinical features of patients infected with 2019 novel coronavirus in Wuhan, China. Lancet. 2020;395(10223):497–506.

9. Pedersen SF, Ho YC. SARS-CoV-2: a storm is raging. J Clin Invest. 2020;130(5):2202–2205.

10. Wang D, Hu B, Hu C, et al. Clinical Characteristics of 138 Hospitalized Patients With 2019 Novel Coronavirus-Infected Pneumonia in Wuhan, China. JAMA. 2020.

11. Mehta P, McAuley DF, Brown M, et al. COVID-19: consider cytokine storm syndromes and immunosuppression. Lancet. 2020;395(10229):1033–1034.

12. Meredith Wadman JC-F, Jocelyn Kaiser, Catherine Matacic. How does coronavirus kill? Clinicians trace a ferocious rampage through the body, from brain to toes. 2020.

13. Vulliamy P, Jacob S, Davenport RA. Acute aorto-iliac and mesenteric arterial thromboses as presenting features of COVID-19. Br J Haematol. 2020.

14. Connors JM, Levy JH. COVID-19 and its implications for thrombosis and anticoagulation. Blood. 2020.

15. Llitjos JF, Leclerc M, Chochois C, et al. High incidence of venous thromboembolic events in anticoagulated severe COVID-19 patients. J Thromb Haemost. 2020.

16. Lodigiani C, Iapichino G, Carenzo L, et al. Venous and arterial thromboembolic complications in COVID-19 patients admitted to an academic hospital in Milan, Italy. Thromb Res. 2020;191:9–14.

17. Al-Samkari H, Karp Leaf RS, Dzik WH, et al. COVID and Coagulation: Bleeding and Thrombotic Manifestations of SARS-CoV2 Infection. Blood. 2020.

18. Tang N, Li D, Wang X, Sun Z. Abnormal coagulation parameters are associated with poor prognosis in patients with novel coronavirus pneumonia. J Thromb Haemost. 2020;18(4):844–847.

19. Bikdeli B, Madhavan MV, Jimenez D, et al. COVID-19 and Thrombotic or Thromboembolic Disease: Implications for Prevention, Antithrombotic Therapy, and Follow-up. J Am Coll Cardiol. 2020.

20. Levi M, Thachil J, Iba T, Levy JH. Coagulation abnormalities and thrombosis in patients with COVID-19. Lancet Haematol. 2020;7(6):e438–e440.

21. Arentz M, Yim E, Klaff L, et al. Characteristics and Outcomes of 21 Critically Ill Patients With COVID-19 in Washington State. JAMA. 2020.

22. Yang X, Yang Q, Wang Y, et al. Thrombocytopenia and its association with mortality in patients with COVID-19. J Thromb Haemost. 2020.

23. Lippi G, Plebani M, Henry BM. Thrombocytopenia is associated with severe coronavirus disease 2019 (COVID-19) infections: A meta-analysis. Clin Chim Acta. 2020;506:145–148.

24. Lippi G, Favaloro EJ. D-dimer is Associated with Severity of Coronavirus Disease 2019: A Pooled Analysis. Thromb Haemost. 2020.

25. Xu P, Zhou Q, Xu J. Mechanism of thrombocytopenia in COVID-19 patients. Ann Hematol. 2020;99(6):1205–1208.

26. Liu Y, Sun W, Guo Y, et al. Association between platelet parameters and mortality in coronavirus disease 2019: Retrospective cohort study. Platelets. 2020;31(4):490–496.

27. Machlus KR, Italiano JE, Jr. The incredible journey: From megakaryocyte development to platelet formation. J Cell Biol. 2013;201(6):785–796.

28. Lefrancais E, Ortiz-Munoz G, Caudrillier A, et al. The lung is a site of platelet biogenesis and a reservoir for haematopoietic progenitors. Nature. 2017;544(7648):105–109.

29. Davi G, Patrono C. Platelet activation and atherothrombosis. N Engl J Med. 2007;357(24):2482–2494.

30. Ridger VC, Boulanger CM, Angelillo-Scherrer A, et al. Microvesicles in vascular homeostasis and diseases. Position Paper of the European Society of Cardiology (ESC) Working Group on Atherosclerosis and Vascular Biology. Thromb Haemost. 2017;117(7):1296–1316.

31. Semple JW, Italiano JE, Freedman J. Platelets and the immune continuum. Nat Rev Immunol. 2011;11(4):264–274.

32. Rick Kapur AZ, Eric Boilard, and John W. Semple. Nouvelle Cuisine: Platelets Served with Inflammation. J Immunol. 2015;12(194).

33. Morrell CN, Aggrey AA, Chapman LM, Modjeski KL. Emerging roles for platelets as immune and inflammatory cells. Blood. 2014;123(18):2759–2767.

34. Sreeramkumar V, Adrover JM, Ballesteros I, et al. Neutrophils scan for activated platelets to initiate inflammation. Science. 2014;346(6214):1234–1238.

35. Imhof BA, Jemelin S, Ballet R, et al. CCN1/CYR61-mediated meticulous patrolling by Ly6Clow monocytes fuels vascular inflammation. Proc Natl Acad Sci U S A. 2016;113(33):E4847–4856.

36. Zuo Y, Yalavarthi S, Shi H, et al. Neutrophil extracellular traps in COVID-19. JCI Insight. 2020.

37. Barnes BJ, Adrover JM, Baxter-Stoltzfus A, et al. Targeting potential drivers of COVID-19: Neutrophil extracellular traps. J Exp Med. 2020;217(6).

38. Martinod K, Wagner DD. Thrombosis: tangled up in NETs. Blood. 2014;123(18):2768–2776.

39. Constantinescu-Bercu A, Grassi L, Frontini M, Salles C, II, Woollard K, Crawley JT. Activated alphaIIbbeta3 on platelets mediates flow-dependent NETosis via SLC44A2. Elife. 2020;9.

40. Lindemann S, Tolley ND, Dixon DA, et al. Activated platelets mediate inflammatory signaling by regulated interleukin 1beta synthesis. J Cell Biol. 2001;154(3):485–490.

41. Karas SP, Rosse WF, Kurlander RJ. Characterization of the IgG-Fc receptor on human platelets. Blood. 1982;60(6):1277–1282.

42. Campbell RA, Schwertz H, Hottz ED, et al. Human megakaryocytes possess intrinsic antiviral immunity through regulated induction of IFITM3. Blood. 2019;133(19):2013–2026.

43. Simon AY, Sutherland MR, Pryzdial EL. Dengue virus binding and replication by platelets. Blood. 2015;126(3):378–385.

44. Koupenova M, Corkrey HA, Vitseva O, et al. The role of platelets in mediating a response to human influenza infection. Nat Commun. 2019;10(1):1780.

45. Le VB, Schneider JG, Boergeling Y, et al. Platelet activation and aggregation promote lung inflammation and influenza virus pathogenesis. Am J Respir Crit Care Med. 2015;191(7):804–819.

46. Cloutier N, Allaeys I, Marcoux G, et al. Platelets release pathogenic serotonin and return to circulation after immune complex-mediated sequestration. Proc Natl Acad Sci U S A. 2018;115(7):E1550–E1559.

47. Boilard E, Pare G, Rousseau M, et al. Influenza virus H1N1 activates platelets through FcgammaRIIA signaling and thrombin generation. Blood. 2014;123(18):2854–2863.

48. Metlay JP, Waterer GW, Long AC, et al. Diagnosis and Treatment of Adults with Community-acquired Pneumonia. An Official Clinical Practice Guideline of the American Thoracic Society and Infectious Diseases Society of America. Am J Respir Crit Care Med. 2019;200(7):e45–e67.

49. Bou Khzam L, Hachem A, Zaid Y, Boulahya R, Mourad W, Merhi Y. Soluble CD40 ligand impairs the anti-platelet function of peripheral blood angiogenic outgrowth cells via increased production of reactive oxygen species. Thromb Haemost. 2013;109(5):940–947.

50. Hachem A, Yacoub D, Zaid Y, Mourad W, Merhi Y. Involvement of nuclear factor kappaB in platelet CD40 signaling. Biochem Biophys Res Commun. 2012;425(1):58–63.

51. Crackower MA, Sarao R, Oudit GY, et al. Angiotensin-converting enzyme 2 is an essential regulator of heart function. Nature. 2002;417(6891):822–828.

52. Danilczyk U, Penninger JM. Angiotensin-converting enzyme II in the heart and the kidney. Circ Res. 2006;98(4):463–471.

53. Chen Y, Chen L, Deng Q, et al. The presence of SARS-CoV-2 RNA in the feces of COVID-19 patients. J Med Virol. 2020.

54. Perry MJ, Arrington S, Neumann LM, Carrell D, Mores CN. It is currently unknown whether SARS-CoV-2 is viable in semen or whether COVID-19 damages sperm. Andrology. 2020.

55. Aird WC. The role of the endothelium in severe sepsis and multiple organ dysfunction syndrome. Blood. 2003;101(10):3765–3777.

56. Dolmatova EV, Wang K, Mandavilli R, Griendling KK. The effects of sepsis on endothelium and clinical implications. Cardiovasc Res. 2020.

57. Denis MM, Tolley ND, Bunting M, et al. Escaping the nuclear confines: signaldependent pre-mRNA splicing in anucleate platelets. Cell. 2005;122(3):379–391.

58. Melki I, Tessandier N, Zufferey A, Boilard E. Platelet microvesicles in health and disease. Platelets. 2017;28(3):214–221.

59. Owens AP, 3rd, Mackman N. Microparticles in hemostasis and thrombosis. Circ Res. 2011;108(10):1284–1297.

60. Zaid Y, Senhaji N, Darif Y, Kojok K, Oudghiri M, Naya A. Distinctive roles of PKC delta isozyme in platelet function. Curr Res Transl Med. 2016;64(3):135–139.

61. Murugappan S, Tuluc F, Dorsam RT, Shankar H, Kunapuli SP. Differential role of protein kinase C delta isoform in agonist-induced dense granule secretion in human platelets. J Biol Chem. 2004;279(4):2360–2367.

62. Buchholz UJ, Bukreyev A, Yang L, et al. Contributions of the structural proteins of severe acute respiratory syndrome coronavirus to protective immunity. Proc Natl Acad Sci U S A. 2004;101(26):9804–9809.

63. Koupenova M, Vitseva O, MacKay CR, et al. Platelet-TLR7 mediates host survival and platelet count during viral infection in the absence of platelet-dependent thrombosis. Blood. 2014;124(5):791–802.

64. Monteil V, Kwon H, Prado P, et al. Inhibition of SARS-CoV-2 Infections in Engineered Human Tissues Using Clinical-Grade Soluble Human ACE2. Cell. 2020;181(4):905–913 e907.

65. Duerschmied D, Suidan GL, Demers M, et al. Platelet serotonin promotes the recruitment of neutrophils to sites of acute inflammation in mice. Blood. 2013;121(6):1008–1015.

66. Andre P, Prasad KS, Denis CV, et al. CD40L stabilizes arterial thrombi by a beta3 integrin--dependent mechanism. Nat Med. 2002;8(3):247–252.

67. Nunez-Avellaneda D, Mosso-Pani MA, Sanchez-Torres LE, Castro-Mussot ME, Corona-de la Pena Na, Salazar MI. Dengue Virus Induces the Release of sCD40L and Changes in Levels of Membranal CD42b and CD40L Molecules in Human Platelets. Viruses. 2018;10(7).

68. Boudreau LH, Duchez AC, Cloutier N, et al. Platelets release mitochondria serving as substrate for bactericidal group IIA-secreted phospholipase A2 to promote inflammation. Blood. 2014;124(14):2173–2183.

69. Maugeri N, Capobianco A, Rovere-Querini P, et al. Platelet microparticles sustain autophagy-associated activation of neutrophils in systemic sclerosis. Sci Transl Med. 2018;10(451).

